# Collaborative forecasting of influenza-like illness in Italy: the Influcast experience

**DOI:** 10.1101/2024.09.09.24313361

**Authors:** Stefania Fiandrino, Andrea Bizzotto, Giorgio Guzzetta, Stefano Merler, Federico Baldo, Eugenio Valdano, Alberto Mateo Urdiales, Antonino Bella, Francesco Celino, Lorenzo Zino, Alessandro Rizzo, Yuhan Li, Nicola Perra, Corrado Gioannini, Paolo Milano, Daniela Paolotti, Marco Quaggiotto, Luca Rossi, Ivan Vismara, Alessandro Vespignani, Nicolò Gozzi

## Abstract

Collaborative hubs that integrate multiple teams to generate ensemble projections and forecasts for shared targets are now regarded as state-of-the-art in epidemic predictive modeling. In this paper, we introduce Influcast, Italy’s first epidemic forecasting hub for influenza-like illness. During the 2023/2024 winter season, Influcast provided 20 rounds of forecasts, involving five teams and eight models to predict influenza-like illness incidence up to four weeks in advance at the national and regional administrative level. The individual forecasts were synthesized into an ensemble and bench-marked against a baseline model. The ensemble forecasts consistently outperformed both individual models and baseline forecasts, demonstrating superior accuracy at national and sub-national levels across various metrics. Despite a decline in absolute performance over longer horizons, the ensemble model outperformed the baseline in all considered time frames. These findings underscore the importance of multimodel forecasting hubs in producing consistent short-term influenza-like illnesses forecasts that can inform public health preparedness and mitigation strategies.

## 1 Introduction

In modern public health, infectious disease forecasting and scenario modeling play a crucial role. These approaches provide stakeholders and policymakers with essential tools to anticipate and respond to infectious diseases by integrating mathematical models, advanced analytics, and data [1–3]. Over the past decade, inspired by best practices in disciplines like meteorology [4], collaborative hubs have emerged as an innovative tool for epidemiological forecasts [5, 6]. More in detail, a forecasting hub is a consortium of several modeling teams that contribute their forecasts to a common set of targets. The outcomes are generally combined into an ensemble forecast (representing the hub consensus), which has proven to be more accurate and consistent in time than each individual contributing forecast [7, 8]. In the context of epidemiological forecasts, this approach was pioneered by the Centers for Disease Control and Prevention with the 2013-2014 Influenza Season Challenge [9] and later expanded to support the response to several health threats including Ebola [10], West Nile Virus [11] Dengue [12], Chikungunya [13], Influenza [14, 15], and more recently, COVID-19 [7, 8, 16]. In various instances, different targets have been considered, ranging from cases to hospitalizations and disease-related mortality.

Here, we present *Influcast*, the first Italian epidemic forecasting hub on influenza-like illness (ILI). Influenza and other respiratory diseases impose a significant burden on healthcare and hospital systems worldwide and constitute one of the primary economic costs for public health systems [17, 18]. Forecasting the impact of respiratory diseases provides a wealth of actionable information that enables the allocation of resources (e.g., hospital beds, medical staff, vaccines), and helps in fine-tuning communication campaigns to raise public awareness and promote preventive measures. In Italy, the epidemiological surveillance of respiratory diseases typically runs every winter season from week 42 to week 17 of the following year. Sentinel doctors diagnose potential cases of ILI based on specific symptoms and report them to the Istituto Superiore di Sanit`a (ISS, the Italian National Institute of Health). This data is then aggregated by the ISS, which releases weekly estimates of ILI incidence at both national and sub-national (19 regions and 2 autonomous provinces) levels [19]. Due to the time required for reporting and aggregation, the provided report pertains to the week preceding its release date.

Influcast was launched in the 2023/2024 winter season to forecast ILI incidence in Italy and its regions up to four weeks ahead. Over 20 forecasting rounds, 5 independent research teams contributed a total of 8 different models. Individual model’s forecasts were combined into an ensemble forecast and evaluated in real-time against a baseline forecasting model which consistently predicts as median value the last data point within the calibration period and whose predictive intervals are estimated on past data. Across various evaluation metrics and spatial units, the ensemble model consistently emerged as the best performer. When assessing aggregated performance metrics, the ensemble model frequently ranks among the top, most often placing in the upper half across different criteria. Although its absolute forecasting accuracy declines over longer horizons, the ensemble consistently outperforms the baseline model at all time frames. At the sub-national level, the ensemble remains the leading model, surpassing the baseline in every region and outperforming individual models in most cases.

Overall, our findings confirm the effectiveness of collaborative forecasting and ensemble models in delivering reliable short-term forecasts of influenza-like illness incidence. The results demonstrate that pooling predictions from multiple modeling teams not only improves accuracy but also increases the robustness of forecasts. These insights underscore the value of multi-model approaches in public health, supporting better-informed decision-making for managing influenza-like illness.

## 2 Results

Influcast generated 20 forecasting rounds during the 2023/24 influenza season, from week 2023-46 to 2024-13. In each round, modeling teams were asked to submit forecasts on ILI incidence in Italy and its regions for the next four weeks. Incidence is calculated as reported cases per 1, 000 patients (i.e., the population covered by the network of sentinel physicians). Only probabilistic forecasts in the form of quantiles were accepted (more details on the submission format are provided in Sec. 4.1). Each week, individual forecasts were combined into an ensemble forecast and evaluated against a baseline model which consistently predicts as median value the last data point within the calibration period and whose prediction intervals are estimated on past data. Details on the calculations for both ensemble and baseline models are provided in Sec. 4.2.

### 2.1 Contributing models

By the end of the season, 5 different modeling teams had submitted 8 individual models, resulting in a total of 200, 000 forecasting points. Of the eight models submitted, four were mechanistic models, two were semi-mechanistic models, and the remaining two were statistical models. Among the four mechanistic models, two adopted a metapopulation approach, while the other two used a single-population approach. Besides historical data for fitting, the models incorporated additional data, including i) mobility data, ii) contact data, iii) population distribution data, and iv) participatory surveillance data. We refer the reader to the Supplementary Information for further details on the models.

Figure 1A shows an example of one-week-ahead ensemble forecasts (50% and 90% prediction intervals) for Italy and its regions in different forecasting rounds. In the Supplementary Information, we show also two, three, and four weeks ahead ensemble forecasts in different spatial units. Figure 1B illustrates the number of models submitted for each round. Despite small fluctuations, the number of submitted models remained fairly stable across rounds, with four out of eight total models submitting forecasts at the sub-national level as well.

**Figure 1.**
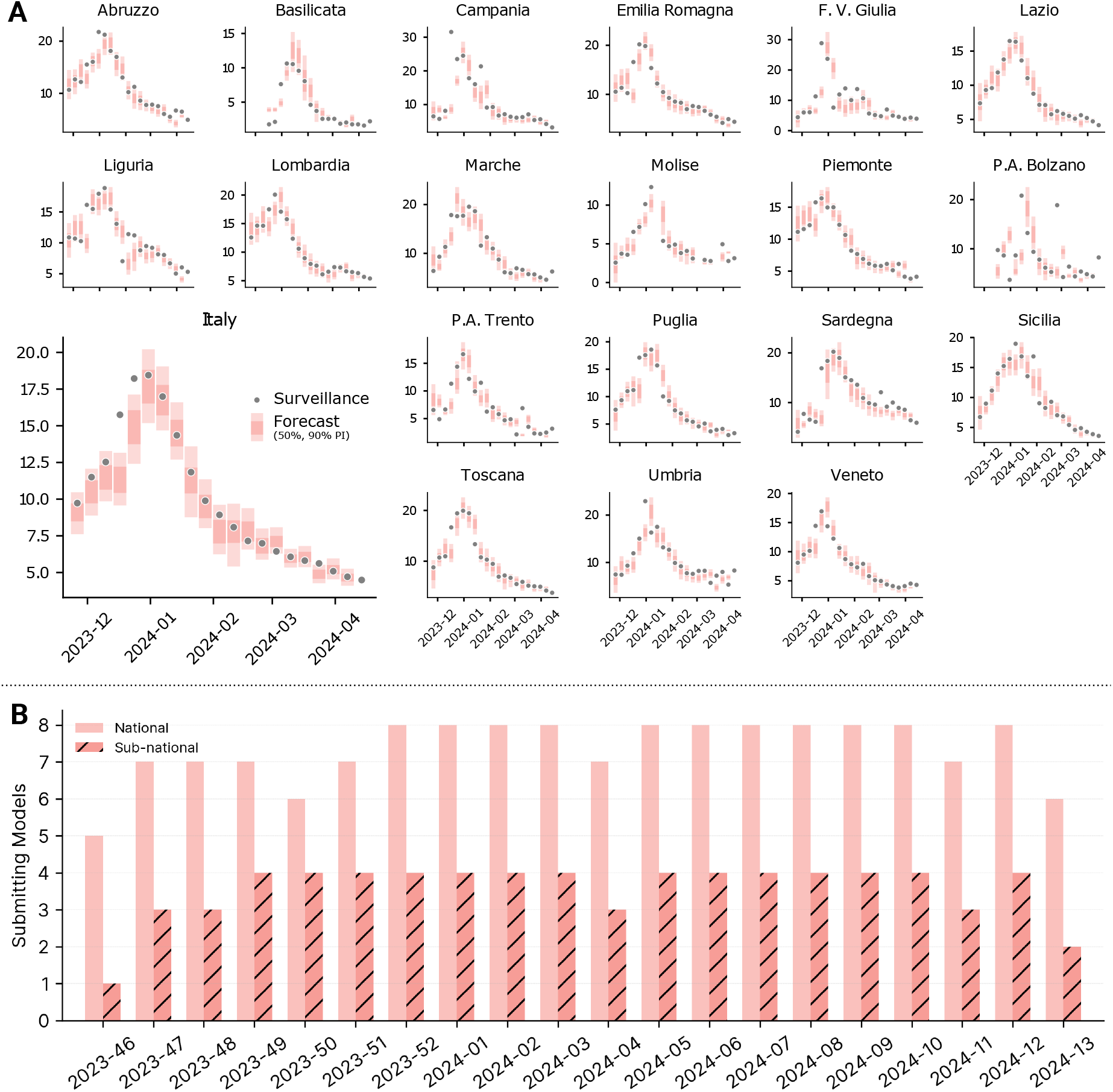
**A)** One-week ahead ensemble forecasts (50% and 90% prediction intervals) at the national and sub-national level in different forecasting rounds. **B)** Number of models submitting forecasts at the national and sub-national level.

### 2.2 Performance of submitted models

Defining what makes a model “good” is challenging, particularly because a model’s performance depends on the target metric considered. In this context, we evaluate models’ predictive performance based on the paradigm *of maximizing the sharpness of the predictive distributions subject to calibration* [20]. In other words, we aim for prediction intervals to be as narrow as possible (and closely aligned with real data), while ensuring that forecasted probabilities match the observed frequencies.

#### National forecasts

Table 1 presents the aggregated performance of individual models using several metrics to evaluate the predicted ILI incidence at the national level. The first metric considered is the relative mean absolute error of the median forecast (relative MAE). The procedure to compute the relative MAE is detailed in Sec. 4.3. In general, aggregating forecasts from all rounds and horizons, each model is compared to the others submitted in the same round in terms of MAE, and the obtained value is then divided by the value for the baseline model. Consequently, values of relative MAE below 1 indicate better performance, while values above 1 indicate worse performance compared to the baseline. Among the models, *Mechanistic-1* shows the lowest relative MAE (0.56), followed closely by *Mechanistic-3* (0.58) and the ensemble (0.59). Notably, a relative MAE of around 0.5 indicates that the model reduces uncertainty in predictions by approximately 50% compared to the baseline. Then we find *Mechanistic-4*, with a relative MAE of 0.73, *Statistical-2* (0.77), *Semi-mechanistic-2* (0.84), and *Statistical-1* (0.99). The remaining two models show a relative MAE greater than one, *Semi-mechanistic-1* (1.80) and *Mechanistic-2* (1.86).

**Table 1.** Models forecasting performance. Performance of different models in terms of relative MAE of the median, relative WIS, 50% and 90% coverage.

| Model | N. of Rounds | Relative MAE | Relative WIS | Coverage (50%) | Coverage (90%) |
| --- | --- | --- | --- | --- | --- |
| *Mechanistic-1* | 19 | **0.56** (1<sup>st</sup>) | **0.54** (1<sup>st</sup>) | 0.75 | **0.89** (1<sup>st</sup>) |
| *Mechanistic-2* | 20 | 1.86 | 1.64 | 0.04 | 0.49 |
| *Mechanistic-3* | 20 | 0.58 (2<sup>nd</sup>) | 0.58 (3<sup>rd</sup>) | 0.47 (3<sup>rd</sup>) | 0.70 (3<sup>rd</sup>) |
| *Mechanistic-4* | 19 | 0.73 | 0.73 | 0.16 | 0.37 |
| *Semi-mechanistic-1* | 16 | 1.80 | 2.14 | 0.06 | 0.27 |
| *Semi-mechanistic-2* | 14 | 0.84 | 1.04 | 0.19 | 0.30 |
| *Statistical-1* | 20 | 0.99 | 1.09 | 0.17 | 0.43 |
| *Statistical-2* | 19 | 0.77 | 0.85 | 0.11 | 0.31 |
| *Ensemble* | 20 | 0.59 (3<sup>rd</sup>) | 0.57 (2<sup>nd</sup>) | **0.51** (1<sup>st</sup>) | 0.80 (2<sup>nd</sup>) |
| *Baseline* | 20 | 1.0 | 1.0 | 0.49 (2<sup>nd</sup>) | 0.66 |

We also consider the relative weighted interval score (relative WIS), a metric that evaluates not only the accuracy of the median but also the accuracy of the prediction intervals in bounding actual data (see Sec. 4.3). The relative WIS is also referenced to the baseline value. In terms of relative WIS, *Mechanistic-1* is the best (0.54), followed by the ensemble (0.57) and *Mechanistic-3* (0.58). In this case, four models have a relative WIS greater than 1.

Finally, to assess calibration of models we evaluate the 90% and 50% coverage. Coverage is defined as the proportion of points falling within the specified intervals (more details in Sec. 4.3). In a well-calibrated model, forecasted probabilities will match the observed frequencies. For example, 50% predictive intervals will contain exactly 50% of observations in a perfectly calibrated model (i.e., the actual coverage is equal to the nominal coverage). The ensemble achieves an almost perfect 50% coverage (0.51), while it is slightly under-confident for 90% coverage (0.80). Among the other models, *Mechanistic-3* follows the ensemble for 50% coverage (0.47), while *Mechanistic-1* performs the best for 90% coverage (0.89). In the Supplementary Information, we present coverage plots for different models and forecasting horizons, demonstrating that the ensemble model maintains good coverage also across other intervals.

We observe that, on average, mechanistic models perform better. For instance, in terms of relative MAE, the top two models are mechanistic. Similarly, when considering WIS and 90% coverage, the top and third-ranked models are mechanistic, with the ensemble model ranking second in both cases. For 50% coverage, the third and fourth models are mechanistic, while the first and second models are the ensemble and the baseline, respectively.

Figure 2 shows the standardized rank for the WIS and the absolute error of the median of different models across all horizons and forecasting rounds. A model showing the best (worst) performance for a given horizon in a given forecasting round will have a standardized score of 1.0 (0.0). More details are provided in Sec. 4.3. When considering the WIS score, we identify roughly three groups of models. *Mechanistic-3*, the ensemble, and *Mechanistic-4* are the top three, with median ranks higher than 0.7 (0.78, 0.75, and 0.71, respectively). Following this, there is a group of models with scores between 0.30 and 0.70. These include *Mechanistic-1* (0.62), *Semi-mechanistic-2* (0.56), *Statistical-2* (0.44), *Statistical-1* (0.33), and the baseline (0.33). Lastly, we have a group of two models with median ranks lower than the baseline (*Semi-mechanistic-1* and *Mechanistic-2*).

**Figure 2.**
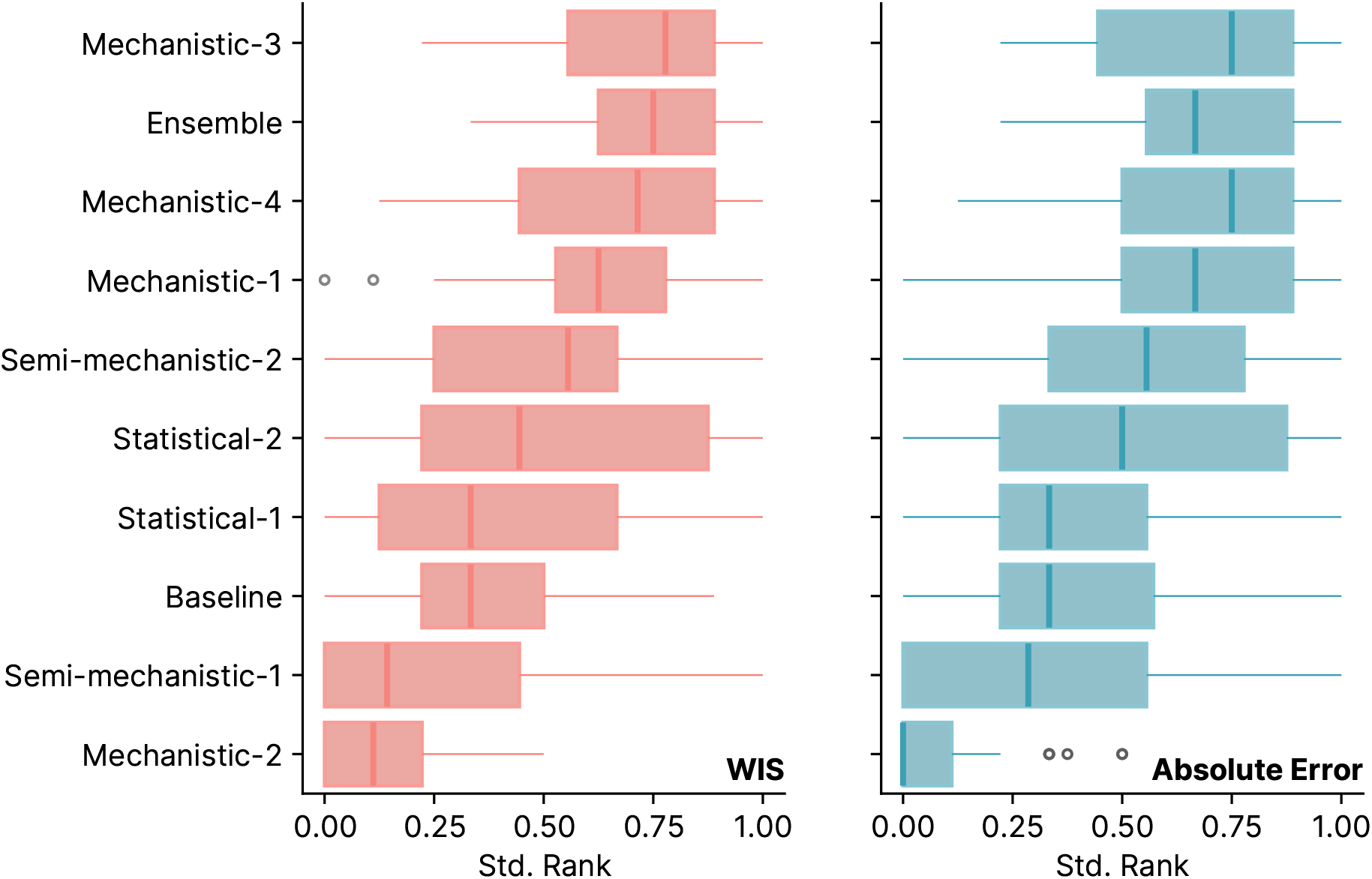
Standardized rank of different forecasting models. Standardized rank plot (WIS and absolute error of the median) for different models computed combining different horizons and forecasting rounds. Boxplots show the distribution of standardized ranks: the box boundaries represent the interquartile range (IQR), the line inside the box indicates the median and the whiskers extend to 1.5 times the IQR from the quartiles. Outliers are shown as individual points.

When considering the absolute error, we see a similar pattern, with the ensemble ranking third. Interestingly, despite not having the highest standardized rank, the ensemble is the model that most frequently appears in the top half across the two metrics. In fact, the ensemble ranks in the top half more than 75% of the time in both cases.

In Fig. 3A, we present the WIS of different models (averaged over a 4-week horizon) divided by the WIS of the baseline across various forecasting rounds. Values below 1 indicate better performance relative to the baseline, while values above 1 indicate worse performance. The performance of the ensemble model is highlighted in red. The background of the plot also displays the evolution of reported ILI incidence. Overall, the ensemble model generally provides significantly better forecasts compared to the baseline, with only a few exceptions. Indeed, we observe a reduction in performance during the forecasting rounds for weeks 49 and 50, just before the epidemic peak. This pattern is observed for the majority of models. This period is typically the most uncertain phase of the epidemic. The reduction in performance can also be attributed to the flattening of the curve around the peak, where the baseline model performs relatively better. Additionally, around the peak, prediction intervals may widen due to increased uncertainty. Interestingly, while the performance of a few models degrades towards the end tail of the epidemic, the ensemble model’s performance remains consistently below 1, indicating sustained superior performance. In the Supplementary Information, we repeat this analysis for the absolute error of the median, finding analogous results.

**Figure 3.**
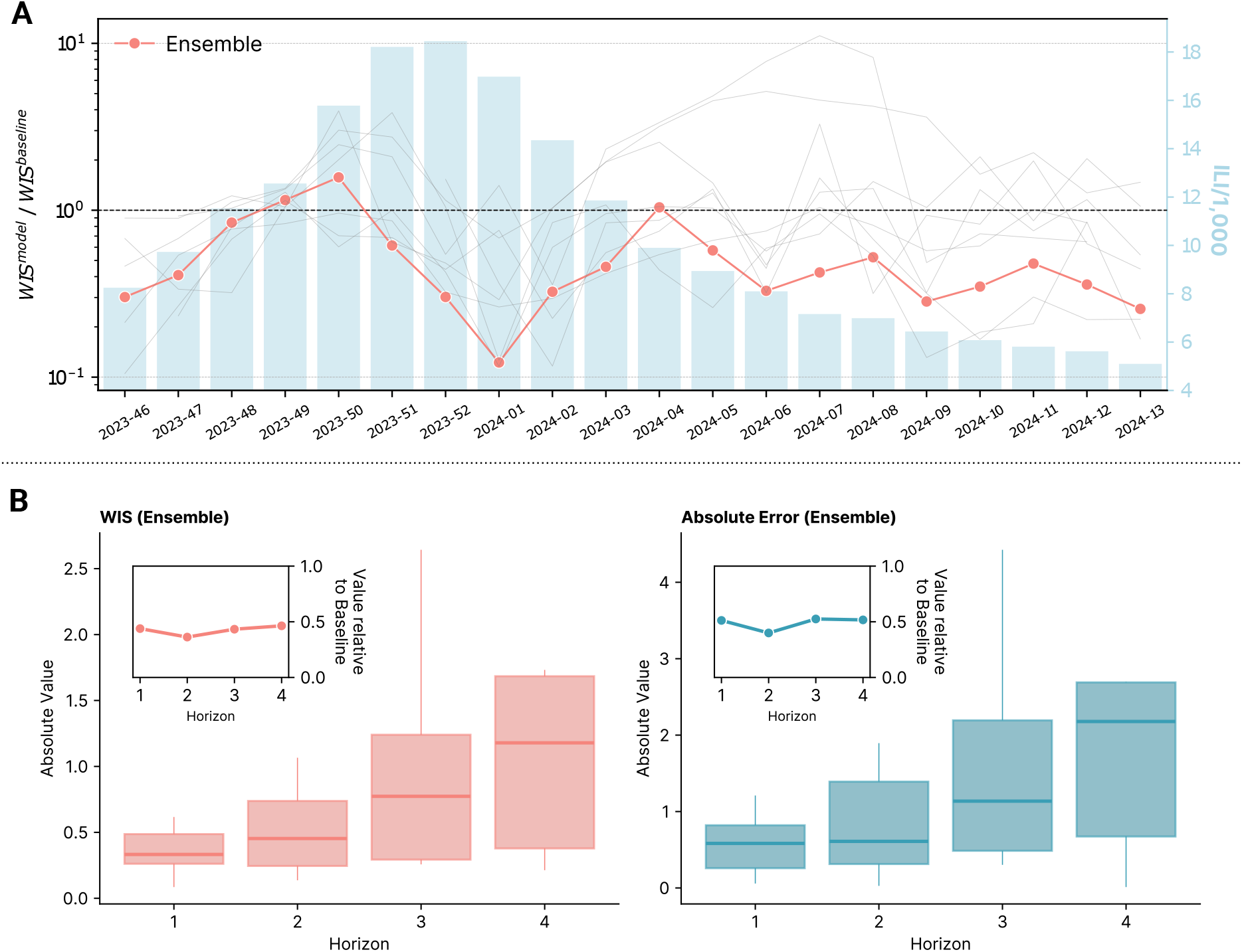
Models performance in time and by horizon. **A)**Comparison of the average WIS of different models to the baseline across different forecast rounds. The ensemble model is highlighted in red. The background displays the reported ILI incidence for the corresponding weeks. **B)** On the left, we show the absolute WIS values of the Ensemble for different forecasting horizons (from 1 to 4 weeks ahead). In the inset, the figure also shows the median WIS relative to the baseline model by horizons. On the right, we repeat the analysis considering the absolute error of the median as a performance metric. The box boundaries represent the interquartile range (IQR), the line inside the box indicates the median and the whiskers extend to 1.5 times the IQR from the quartiles.

In Fig. 3B, we focus on the ensemble and show its performance in terms of absolute WIS and absolute error of the median for different horizons, from one to four weeks ahead, aggregating results from different rounds. As expected, forecasting performance deteriorates for longer horizons. For WIS, the median value increases from 0.33 for one-week-ahead forecasts to 1.18 for four-week-ahead forecasts, a 3.5-fold increase. For absolute error, the median rises from 0.58 to 2.18, a 3.7-fold increase. In the inset,

the figures also show the WIS and absolute error of the ensemble divided by the WIS and the absolute error of the baseline (median values) for different horizons. In this case, we observe a relatively stable trend, with the ensemble’s performance remaining between 0.3 and 0.5 times the baseline performance across different horizons.

#### Sub-national forecasts

In Figure 4A, we present the forecasting performance at the sub-national level. Two regions (Valle d’Aosta and Calabria) did not activate epidemiological surveillance for the 2023/2024 winter season and therefore were excluded from the analysis. On the left, we show the relative WIS for different regions for models providing forecasts at the sub-national level. The models are ordered from best to worst in terms of performance. The ensemble is the best model, with a relative WIS of less than one in all regions. Regions are ordered from best to worst performance. We notice significant heterogeneity in forecasting performance across regions, with Sicilia, Marche, and Lazio showing the best performance, while Liguria, Provincia Autonoma di Bolzano, and Friuli Venezia Giulia showing the worst performance. On the right, we show the standardized rank of the relative WIS for different models across all regions. The ensemble is the top performer with a median rank of 0.8, while the baseline is the worst in terms of median standardized rank. In the Supplementary Information, we repeat the analyses considering the relative absolute error, finding analogous results.

**Figure 4.**
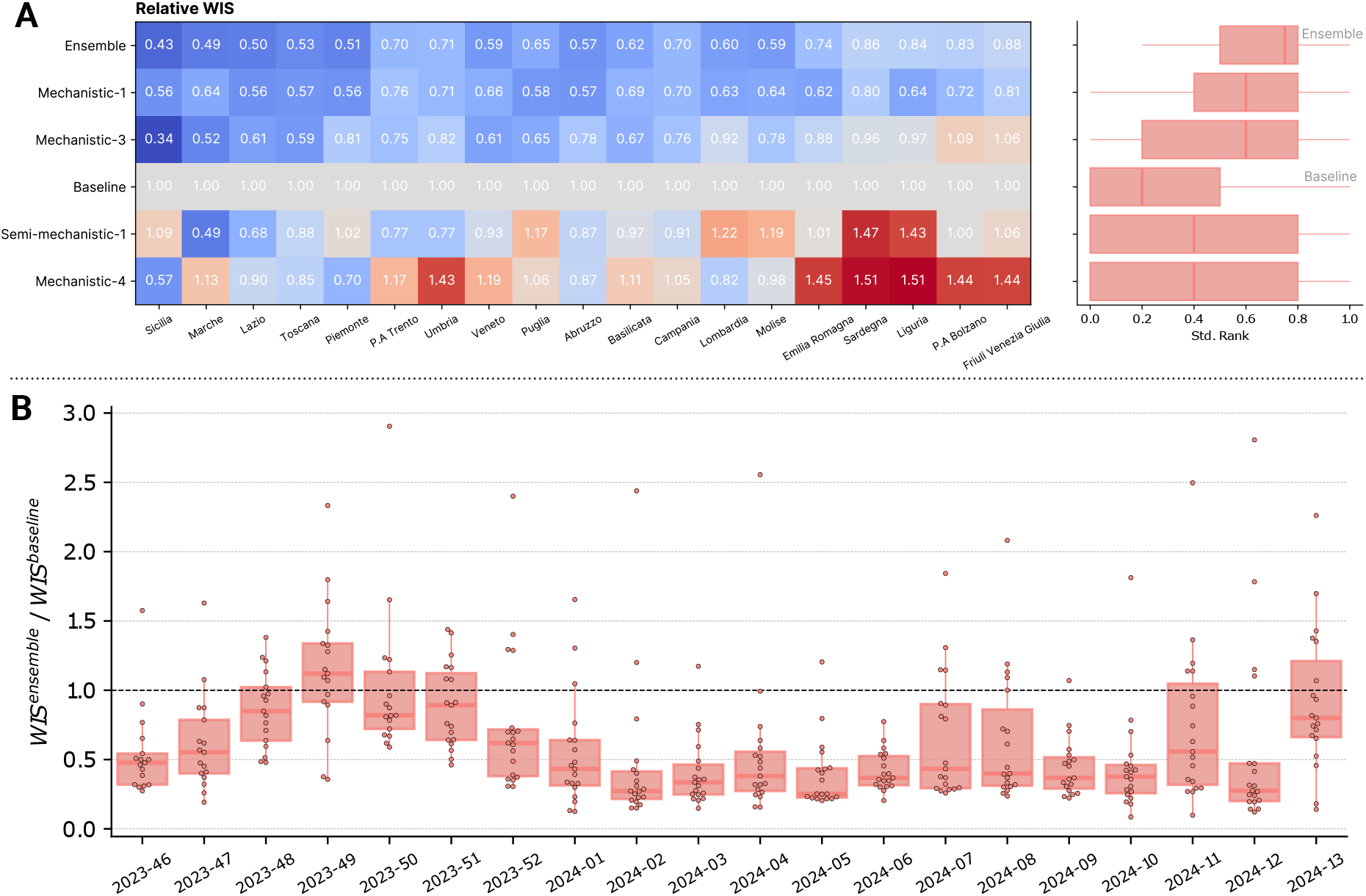
Models subnational forecasting performance. **A)** On the left is shown relative WIS of different models providing sub-national forecasts in different regions. On the right is shown the standardized WIS rank of different models over all regions. The box boundaries represent the interquartile range (IQR), the line inside the box indicates the median and the whiskers extend to 1.5 times the IQR from the quartiles. **B)** Comparison of the average WIS of the ensemble to the baseline across different forecast rounds and regions. The box boundaries represent the interquartile range (IQR), the line inside the box indicates the median and the whiskers extend to 1.5 times the IQR from the quartiles. Individual points are also shown above boxplots.

Finally, in Fig. 4B, we show the performance of the ensemble model over time also in the case of sub-national forecasts. Specifically, boxplots and swarmplots are used to display the average ratio of the WIS of the ensemble model to the baseline model for each forecasting round across different regions. A similar pattern to the national forecasts is observed, with performance decreasing before the epidemic peaks, which occurred between week 2023-50 and week 2024-02 for all regions. Nonetheless, the WIS of the ensemble model is worse than that of the baseline model, in median terms across all regions, only in week 2023-49 throughout the entire period considered. Additionally, also for sub-national forecasts we observe lower performance in the epidemic tail. However, the median performance of the ensemble model across all regions consistently remains below the baseline threshold.

## 3 Discussion

In this paper, we presented Influcast, the first Italian forecasting hub focused on real-time short-term predictions of influenza-like illness incidence. Our performance analysis reveals that: i) most submitted models demonstrated superior performance compared to the baseline across various metrics, and ii) the ensemble model consistently outperforms individual models. Indeed, when considering aggregated performance metrics such as relative WIS, relative MAE, and coverage, the ensemble model is always among the top three performers. Additionally, in terms of standardized rank for both WIS and absolute error, the ensemble consistently ranks in the top three and most frequently appears in the top half for both metrics.

We observed that the forecasting performance of the ensemble deteriorates as the forecast horizon increases, with four-week-ahead predictions being generally more than three times worse than one-week-ahead predictions. Nonetheless, the ensemble consistently performs better than the baseline across all horizons.

When evaluating model performance at the regional level, the ensemble is again the top performer, outperforming the baseline in all regions and each individual model in the majority of cases. Overall, we observed heterogeneous performance across different regions, with some consistently showing better or poorer results compared to others. Although a single round of forecasting limits our ability to explore these differences in depth, we expect that with data from future seasons, we will gain a clearer understanding of the factors driving regional performance, such as data quality or inherent regional characteristics.

We observed that mechanistic models tended to perform better compared to other model classes. Interestingly, past research on influenza forecasts in the US found that statistical models performed similarly or slightly better than mechanistic ones [15], while in the context of COVID-19, top-performing models included both mechanistic and statistical components [8]. However, due to the limited number of models submitted over a single season, this comparison cannot be considered definitive and may be specific to the particular problem being analyzed.

The present work comes with limitations. First, while ILI incidence is a good proxy for the burden of respiratory diseases, it is not a disease-specific signal. Since it is based purely on symptoms, multiple pathogens contribute to this signal, including influenza viruses, coronaviruses (including SARS-CoV-2), and respiratory syncytial viruses. This limits the ability to break down the impact by disease and provides limited information on what is driving ILI dynamics. In future seasons, we aim to overcome these limitations by integrating new data to improve the definition of the forecasting targets, specifically incorporating data from virological surveillance. The Italian National Institute of Health conducts virological surveillance alongside epidemiological surveillance every winter. By combining data from these two surveillance systems, we aim to define targets that are more specific to individual diseases, thereby providing more interpretable information. It is plausible that making targets more disease-specific could enhance the accuracy of forecasts [21]. Second, the limited number of submitted models challenges the comparison of different model classes and the exploration of methodologies to further enhance the ensemble (e.g., weighted or trained ensembles) [22]. However, the number of submitted models meets the minimum requirement to create an effective ensemble, as demonstrated also by a recent study on optimizing the number of ensemble members [23].

Overall, we confirmed the relevance of multi-model efforts in real-time short-term forecasting of infectious diseases, applying for the first time these approaches in Italy for influenza-like illness. More broadly, Influcast is one of the first epidemic forecasting hubs built post-COVID-19 for other respiratory diseases in Europe. We hope it will serve as a model for implementing similar approaches in other countries. Besides providing short-term forecasts during the winter season to support response to respiratory disease epidemics, Influcast overarching goal is to improve outbreak preparedness by building a network of epidemic modelers and public health officials who share best practices and data. These efforts contribute to the establishment of robust collaboration pipelines that can be rapidly deployed in emergency situations.

## 4 Materials and methods

### 4.1 Forecast submission and visualization platform

The infrastructure supporting Influcast consists of two primary components: the GitHub collaborative platform, which is utilized for the submission of forecasts, and a dedicated website for visualizing the output results [24].

During the 2023/24 influenza season, participating teams submitted probabilistic forecasts on ILI incidence for the upcoming four weeks on a weekly basis. Specifically, Influcast considers the number of cases per 1, 000 patients reported by the network of sentinel physicians across Italy as the target variable for the forecasts. This data is communicated every Friday by the Italian National Institute of Health through the RespiVirNet bulletin [19]. The incidence data published pertains to the previous week, while Influcast is updated the following Tuesday, allowing teams time to process the new data and produce new forecasts. Consequently, the forecasts published on Influcast on Tuesday refer to the previous week (for which there is no consolidated public data yet), the current week, and the next two weeks. Detailed information about the forecast format and the submission pipeline is provided on the

Influcast GitHub Wiki page [25]. The Influcast GitHub repository also collects weekly surveillance data, including historical records from past seasons, for both Italy and its regions. In each round, forecasts from each model are automatically combined to produce ensemble predictions, which are stored in the repository as an additional model, alongside the baseline model (more details in Sec. 4.2).

The Influcast web platform features an interface allowing one to examine both individual model forecasts and the ensemble output. An example of the interface is presented in Fig. 5. Forecasts for the next four weeks from each model are represented using prediction intervals. The default values include the 90% and 50% intervals, but users can select a specific prediction interval via the menu located in the bottom right corner of the visualization. By default, the most recent forecasts are displayed. However, users can also explore forecasts from previous weeks by dragging the vertical bar on the graph. White points represent historical data that were unavailable at the time of model calibration. This approach allows for a visual assessment of each model’s predictive ability, enabling verification of whether the actual data fell within the estimated prediction intervals. Through the menu on the left, users can select the geographical unit of interest, either Italy or one of its regions, as well as a specific season. Additionally, it is possible to display the predictions of multiple models by selecting them from the menu located in the bottom left corner.

**Figure 5.**
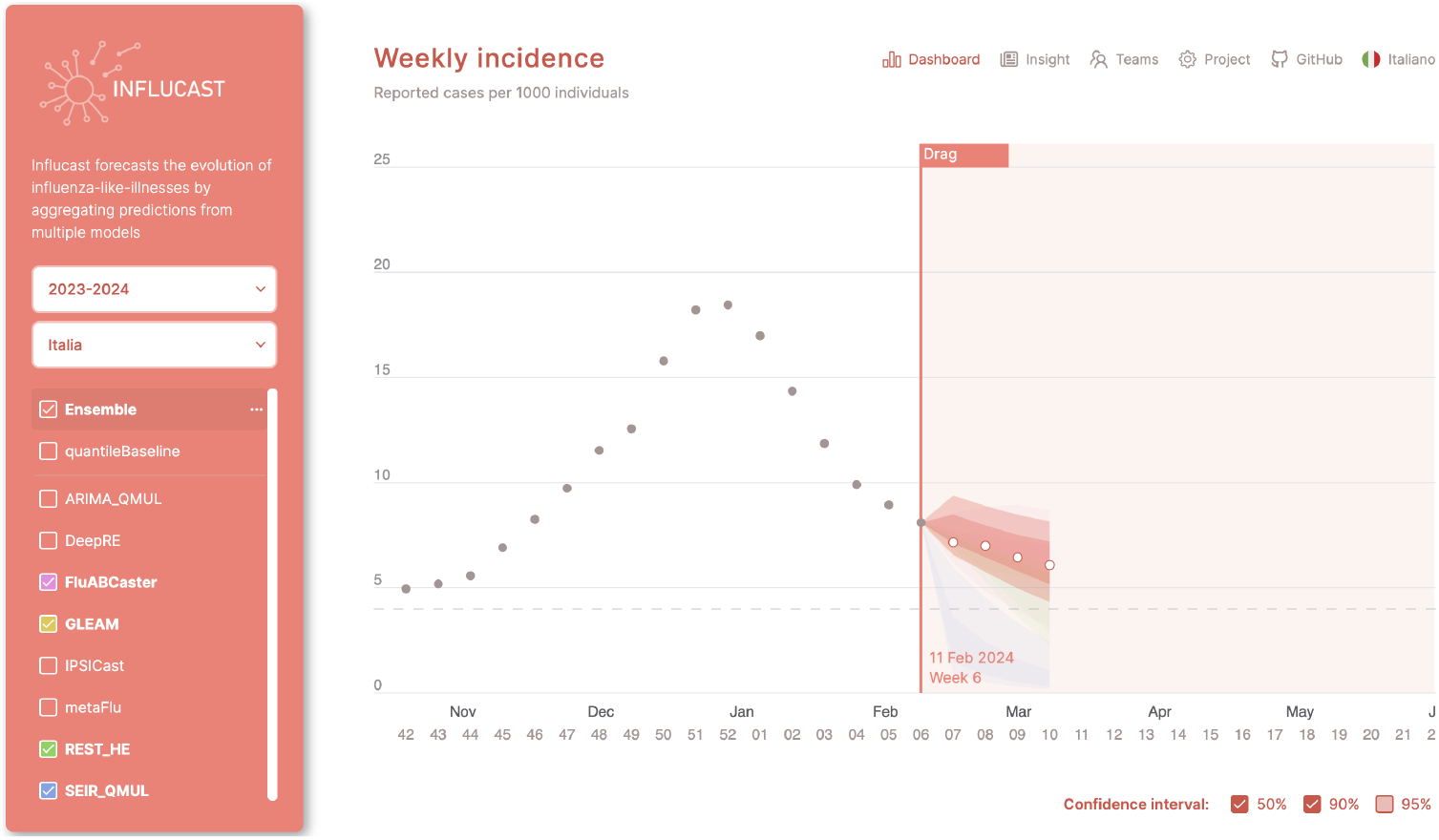
The Influcast web interface. The visualization dashboard allows displaying the four weeks ahead prediction for the ensemble and for specific models, choosing the season, the location and the prediction intervals to show.

### 4.2 Baseline and ensemble models

#### Baseline model

The baseline forecasting model consistently predicts a median value matching the last observed data point during the calibration period. To generate quantiles, we use the previous 1-step increments. Specifically, we compute 1-step differences up to time *t*: *δ* = (*d*_2_, *d*_3_, …, *d*_*t*_). To ensure the median forecast matches the last calibration point, we create a symmetrized set of increments *d*^′^ = (*d, −d*). For a forecast with a maximum horizon of *H*, we sample *H* increments from *δ*^′^. The prediction at horizon *h* is calculated as: 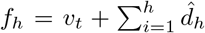 where *v*_*t*_ is the last observed data point. Using this approach, we generate 10, 000 baseline stochastic trajectories to compute quantiles and prediction intervals. We note that this baseline model is the same one used in other epidemiological forecasting challenges, such as FluSight and the US and European COVID-19 Forecasting Hubs [7, 8, 26].

#### Ensemble model

Individual forecasts are combined into an ensemble forecast by taking a simple average across quantiles. This method is also known as quantile averaging or Vincent method [27]. In practice, quantile of level *q* of the ensemble forecast at time *t* will be:

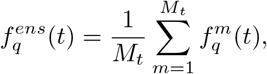

where *M*_*t*_ is the number of models that submitted a forecast at time *t*. All models except for the baseline are included in the ensemble calculation.

### 4.3 Evaluation metrics

#### Weighted Interval Score

The Weighted Interval Score (WIS) is an approximation of the continuous ranked probability score (CRPS) [28]. For a given prediction interval (1 *− α*) *×* 100% (e.g., 90% for *α* = 0.1) of a model’s forecast *F*, the interval score is defined as:

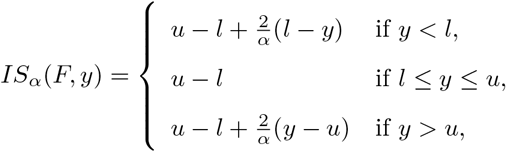

where *u* (*l*) represents the upper (lower) limit of the prediction interval and *y* is the actual observed value. The WIS generalizes the interval score to multiple prediction intervals and is defined as:

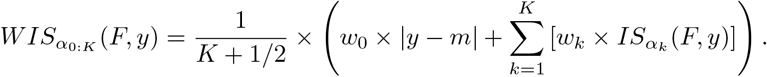

Here, *K* denotes the total number of intervals considered, *m* is the forecast median, and *w*_*k*_ are the non-negative weights assigned to the different intervals. Following a standard approach, we set *w*_0_ = 1*/*2, *w*_*k*_ = *α*_*k*_*/*2, and consider 11 prediction intervals (*α*_*k*_ = 0.02, 0.05, 0.10, 0.20, 0.30, 0.40, 0.50, 0.60, 0.70, 0.80, 0.90).

#### Absolute Error

Given a median forecast *f*_*i*_ and an actual corresponding value *a*_*i*_, the absolute error is simply computed as:

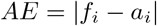

When averaged over different time steps *i* we obtain the mean absolute error.

#### Coverage

Prediction interval coverage is the fraction of times a prediction interval of a given level covers the observed data. Intuitively, 90% coverage is the fraction of times actual data are included in the 90% prediction interval. For a perfectly calibrated model, prediction interval coverage will be equal to the nominal coverage. Following the example, this means that exactly 90% of times points will be included in the 90% prediction interval. Mathematically, for a prediction interval of level *α* and a set of *N* data points *y*_*i*_ it is computed as:

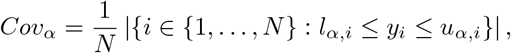

where *l*_*α,i*_ and *u*_*α,i*_ are the lower and upper limits of the prediction interval.

#### Relative performance

We apply the procedure presented in Ref. [8] to compute relative performance. For simplicity, we will present the steps to compute the relative MAE, but an analogous procedure can be performed to obtain the relative WIS. First, for each pair of model *m* and *m*^′^, we compute the pairwise relative MAE skill as:

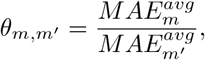

where *MAE*^*avg*^ is the average MAE of model *m* computed using all available forecasts where both *m* and *m*^′^ participated. Then, for each model *m* we compute the geometric mean of different *θ*_*m,m*_*′* :

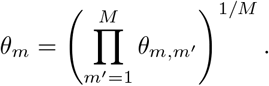

Finally, for easier interpretation, we divide each *θ*_*m*_ by the result of the baseline model:

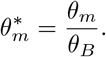

Then, the obtained quantity 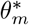 represents the relative MAE of model *m*, adjusted for the difficulty of the forecasts it produced and rescaled such that the baseline model has a relative performance of 1.

It follows that,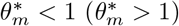 indicates that model *m* is better (worse) than the baseline.**Standardized rank**.

For model *m*, and observation *i* we compute its standardized rank as:

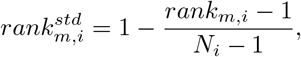

where *N*_*i*_ is the number of models submitting forecasts for observation *i* and *rank*_*m,i*_ is the rank of model *m* and observation *i* according to the metric considered (WIS or absolute error). The model with lowest WIS (or absolute error) for observation *i* will rank first (i.e., *rank*_*m,i*_ = 1), therefore it will have a standardized score of 1. On the contrary, the model with the highest WIS will rank last (*rank*_*m,i*_ = *N*_*i*_) and therefore will have a standardized score of 0.

Evaluation metrics were computed using the *scoringutils* package [29].

### 4.4 Epidemiological data

We use data provided by the Italian National Institute of Health through their weekly bulletins [19]. In Italy, influenza epidemiological surveillance generally runs from week 42 to week 17 each year. Sentinel doctors diagnose potential cases of influenza-like illness based on specific symptoms and report these cases to the National Institute of Health. This data is then aggregated by the institute, which releases weekly estimates of ILI incidence at both national and regional levels. The data typically reflects the previous week; thus, the report published in week *t* contains data from week *t −* 1. Weeks are defined from Monday to Sunday, and the data may be revised due to delays in submissions from some sentinel physicians. Historical incidence data are published on GitHub in a machine-readable format [30].

## Data Availability

All data produced are available online at https://github.com/Predizioni-Epidemiologiche-Italia/Influcast

https://influcast.org/

## Acknowledgements

S.F. acknowledges support from EU grant 101045989 VERDI. E.V. acknowledges support from EU Horizon Europe grant SIESTA (HORIZON-INFRA-2023-EOSC-01-101131957). E.V. and F.B. acknowledge the CINECA award under the ISCRA initiative, for the availability of high-performance computing resources and support. All authors thank Francesco Branda and Luca Cozzuto for providing open access, machine-readable data on influenza-like illness from past seasons.

## Notes

### Competing Interest Statement

The authors have declared no competing interest.

