## Supplementary Information for "Collaborative forecasting of influenza-like illness in Italy: the Influcast experience"

#### Contents

**1 Submitting Models Description** **10**

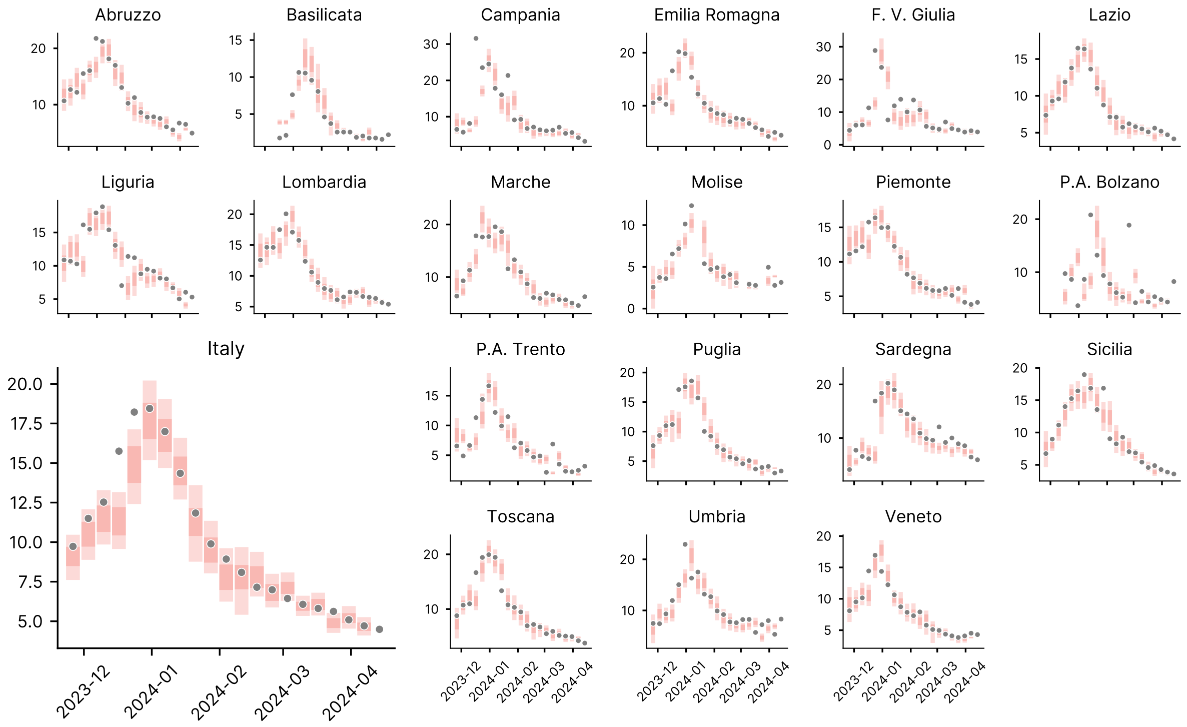

Figure 1: **One-week ahead ensemble forecasts at the national and sub-national level.**

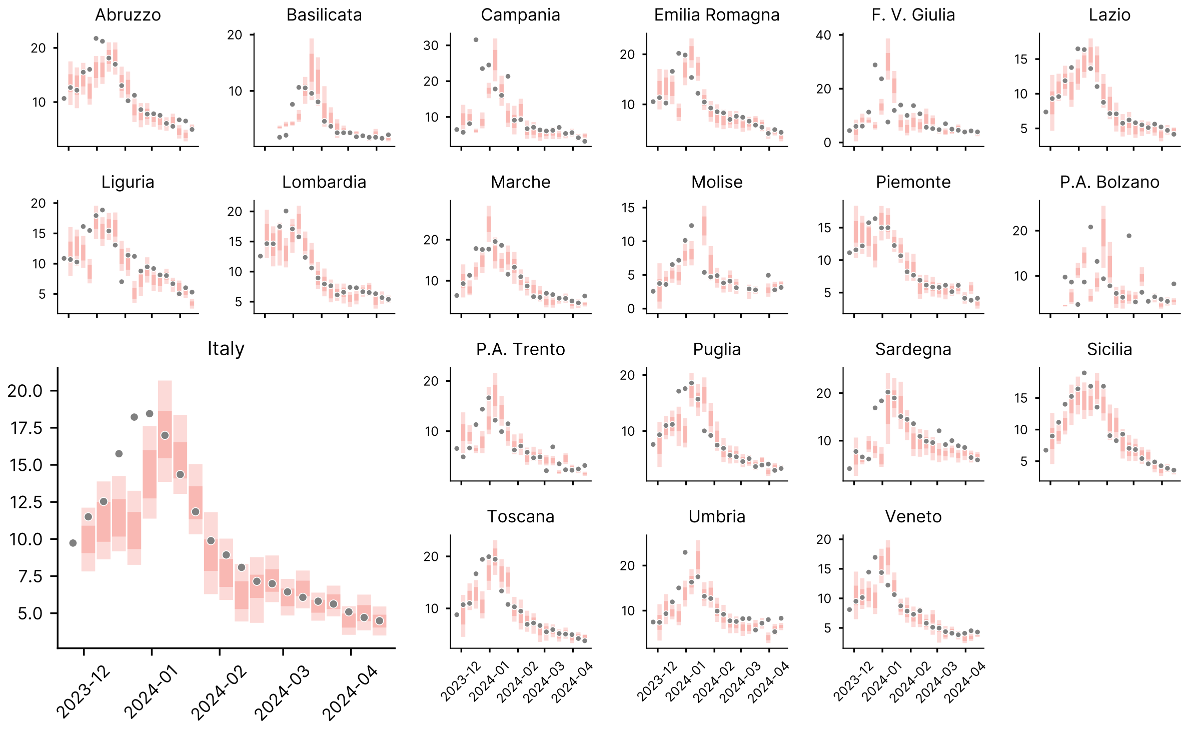

Figure 2: **Two-weeks ahead ensemble forecasts at the national and sub-national level.**

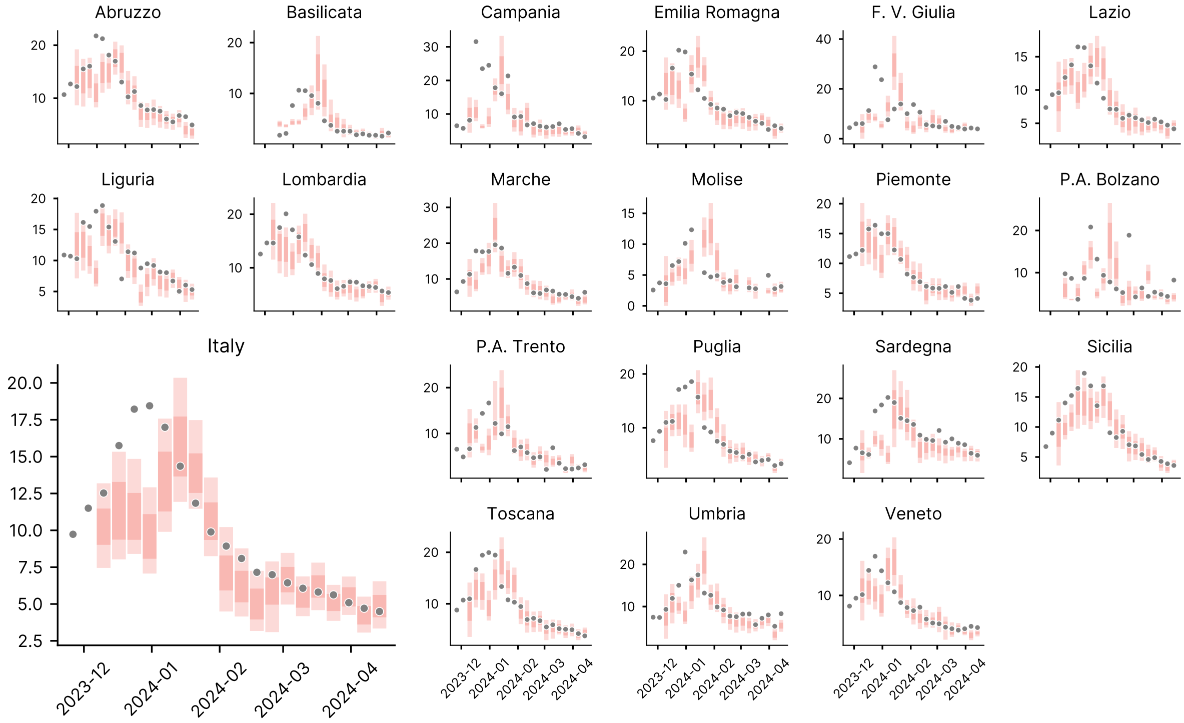

Figure 3: **Three-weeks ahead ensemble forecasts at the national and sub-national level.**

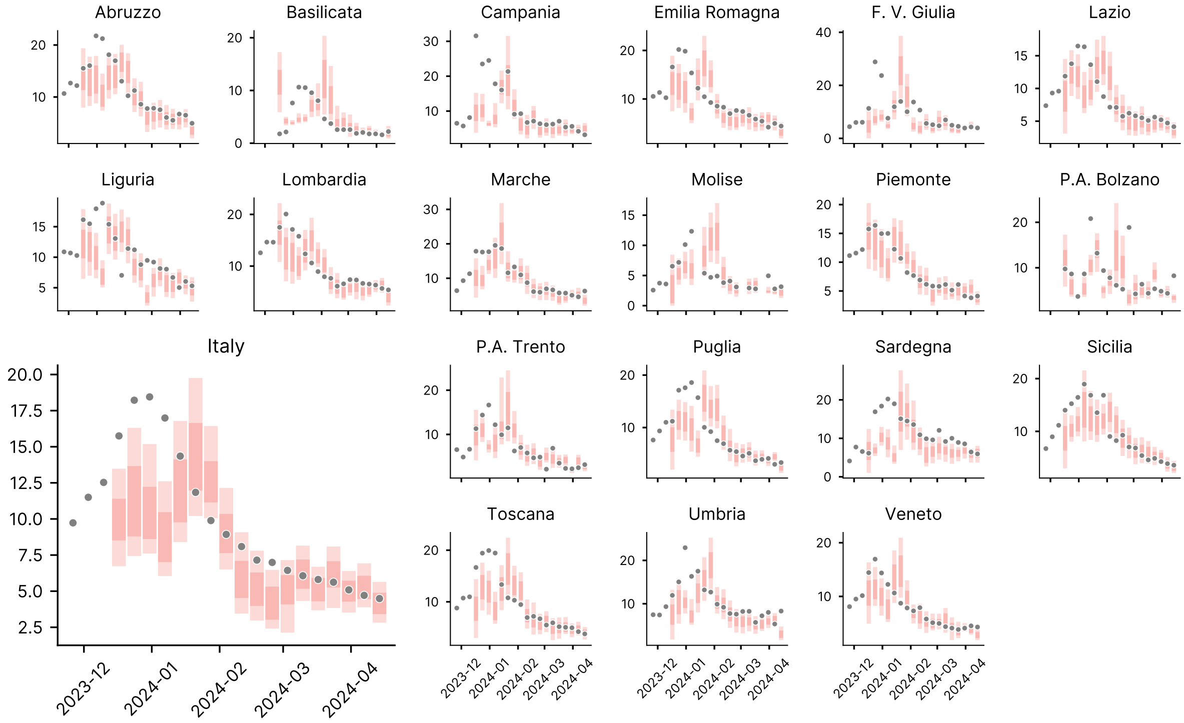

Figure 4: **Four-weeks ahead ensemble forecasts at the national and sub-national level.**

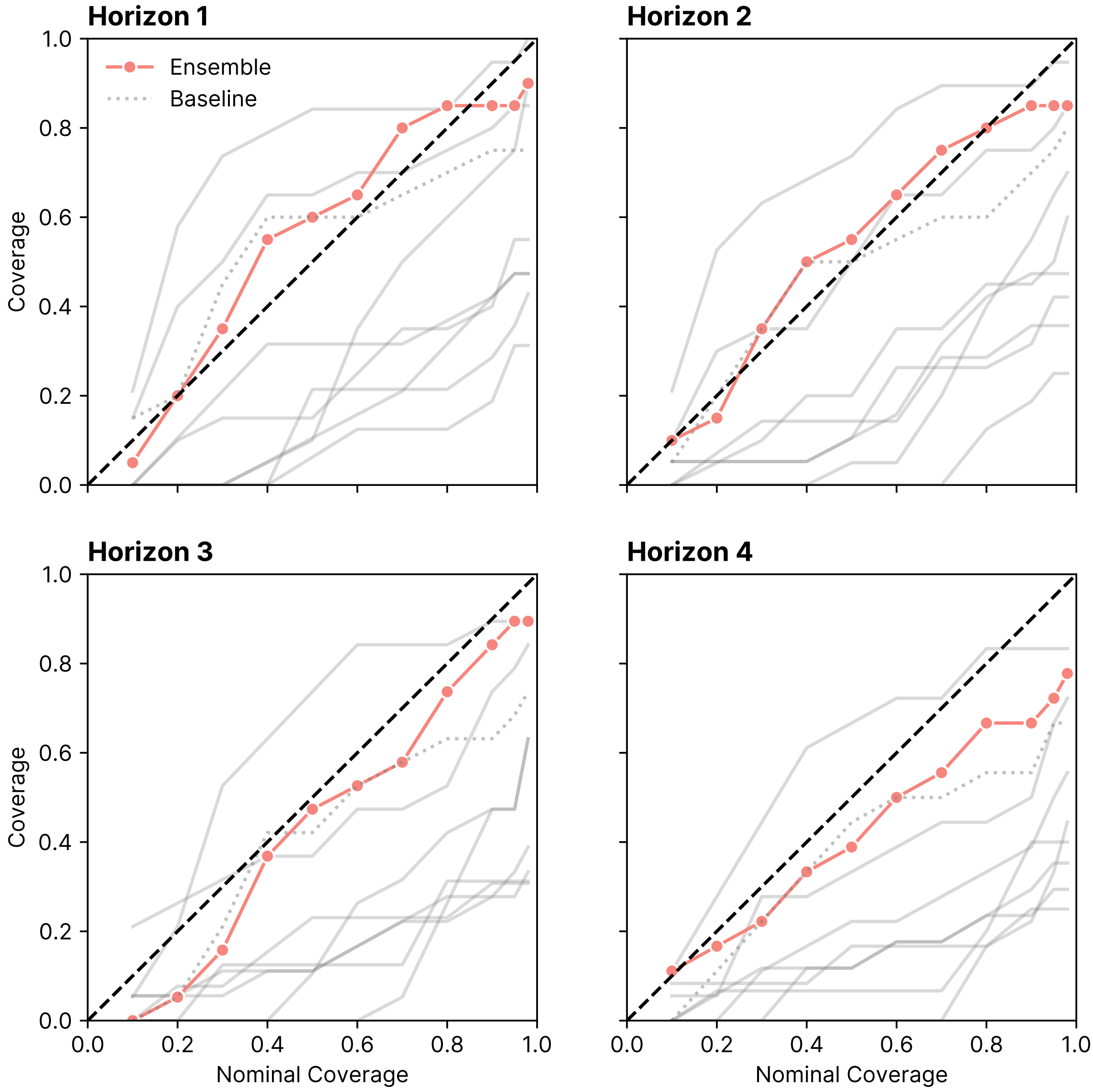

Figure 5: **Coverage by horizons**. We show nominal versus empirical coverage for different models and forecasting horizons. The ensemble is highlighted in red while the baseline as a dotted line.

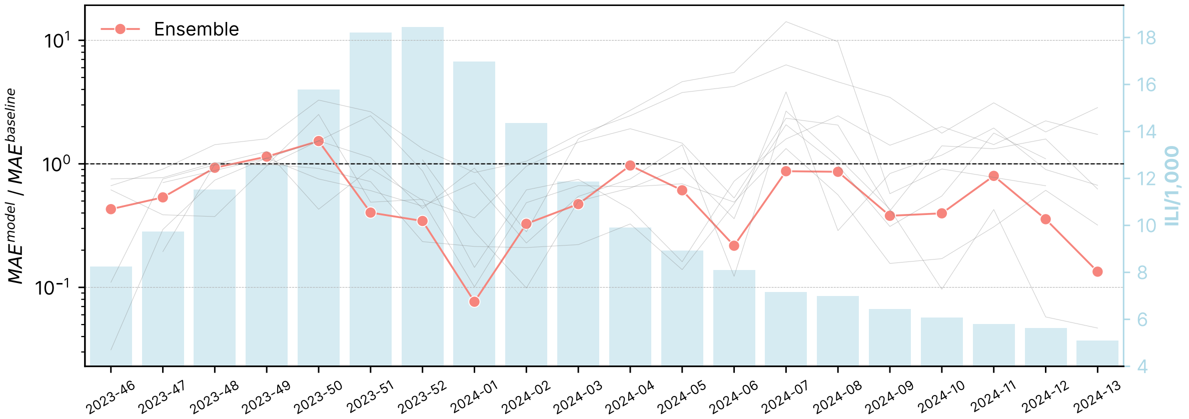

Figure 6: **Models performance in time (Absolute Error)**. **A)** Comparison of the average absolute error of the median of different models to the baseline across different forecast rounds. The ensemble model is highlighted in red. The background displays the reported ILI incidence for the corresponding weeks.

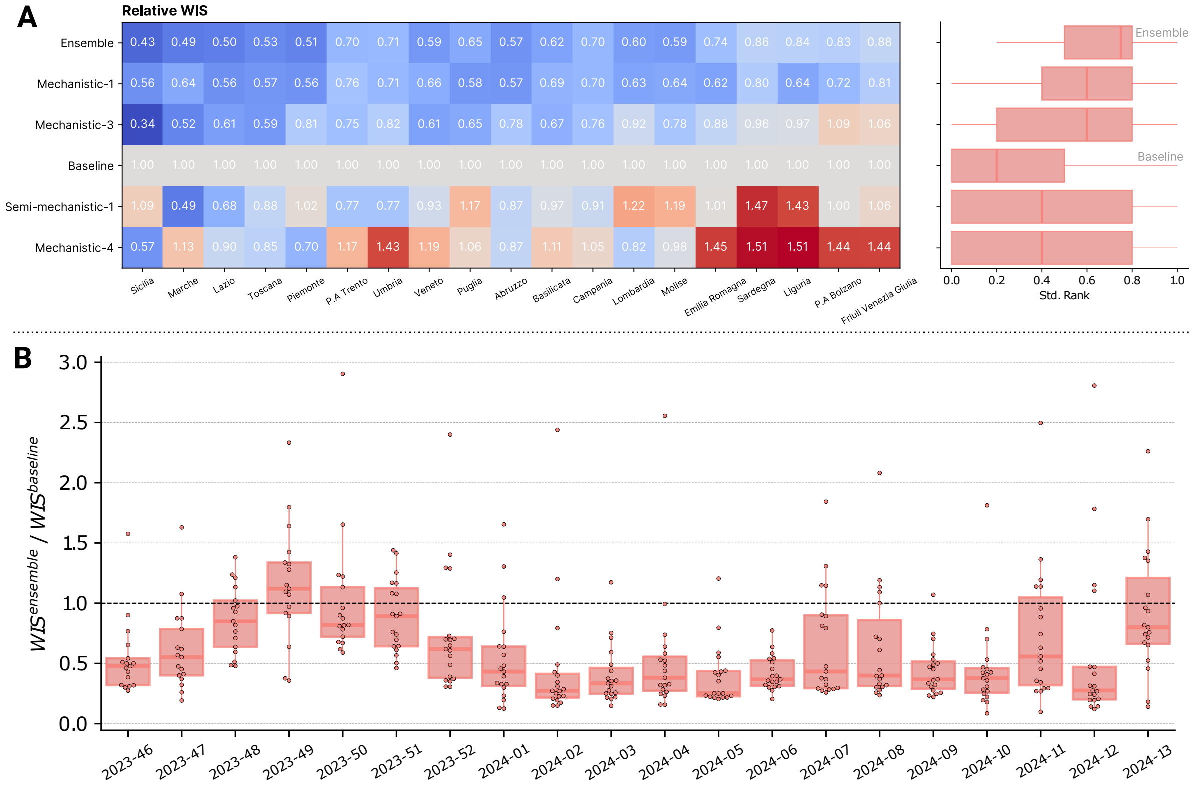

Figure 7: **Sub-national Models Performance (absolute error)**. **A)** On the left is shown the relative AE of different models providing sub-national forecasts in different regions. On the right is shown the standardized AE rank of different models over all regions. **B)** Comparison of the average MAE of the ensemble to the baseline across different forecast rounds and regions.

### 1 Submitting Models Description

We provide here a description of submitting models. Models are categorized based on their category, more in detail we divide them into i) **mechanistic models** explicitly modeling underlying biological processes that govern the spread of diseases (e.g., compartmental models); ii) **semi-mechanistic models** incorporating some mechanistic elements but also statistical components (e.g., non-parametric time-varying transmission rates); iii) **statistical models** not relying on underlying biological mechanisms but instead using statistical techniques to identify patterns and make predictions based on observed data (e.g., time-series forecasting models).

***Mechanistic-1***. Province-based metapopulation model built starting from the model used for COVID-19 in Ref. [1]. The model has been extended to incorporate vaccination. A distribution for the model parameters related to the disease progression has been obtained by calibrating the model to past year influenza seasons and adjusted using available data on the 2023/24 influenza season. To generate the predictions, we simulated 100 realizations of the dynamical system, each one with an independent realization of the model parameters, sampled from the distributions obtained in the calibration process.

***Mechanistic-2***. Age-structured SEIR model calibrated using a simple Approximate Bayesian Computation (ABC) method where the top 1% (out of $200,000$ simulations) are accepted. We used the weighted mean absolute percentage error (wMAPE) as the target metric for calibration.

***Mechanistic-3***. Stochastic, age-structured compartmental model considering a SEIR compartmentalization setup. The population is stratified into 10 age groups ($0-9$, $10-19$, $20-24$, $25-29$, $30-39$, $\ldots$, $70-79$, $80+$) and contacts between different age groups are described by a synthetic contact matrix from Ref. [2]. The model also considers seasonality terms modulating the force of infection. The model is implemented in discrete time (the simulation step is 1 day) and the number of individuals transitioning among compartments is simulated as chain binomial processes. Model calibration is conducted using an Approximate Bayesian Computation technique [3]. Free parameters include transmissibility rate, reporting fraction, and initial immunity.

***Mechanistic-4***. Stochastic, age-structured compartmental model based on a metapopulation approach for simulating the spatiotemporal evolution of the spreading of infectious diseases on a global scale [4, 5]. It uses datasets derived from real-world data about worldwide population density and demographic structure, flight networks and passenger distribution, daily commuting flows, and age-structured contact patterns. The model considers 16 age groups and related contact matrices corresponding to four different settings. The epidemic dynamics within each subpopulation are defined by describing the compartmental structure of a given disease along with the corresponding transition rates and other relevant parameters.

***Semi-mechanistic-1***. Deep Renewal Equation is a model coupling Renewal Theory and Deep Learning. The intuition is that renewal equations in the context of epidemic forecasting are characterized by 3 components: the reproductive factor, the historical series of infected, and the generation time distribution. Putting aside the historical data, which can be assumed to be known, we approximate the other two components of the equation through deep neural networks, allowing for data-driven approximation of these components. The model can be used in two ways: on the one hand, we can predict the epidemic trend, namely the number of infected in time; while on the other hand, the approximated reproductive factor and the generation time distribution can be used to better understand the epidemic itself.

***Semi-mechanistic-2*.** The model is based on the application of the Renewal equation and assumes a distribution of the generation time that is intermediate between those of SARS-CoV-2 and seasonal flu. The final projections are the ensemble of projections based on two different assumptions: i) the reproduction number remains constant at a fixed value (corresponding to the last available estimate) throughout the projection horizon; ii) the reproduction number decreases over time from the last available estimate, with the same trend observed in data from 2003/04 to 2022/23, excluding outlier seasons due to the H1N1 and COVID-19 pandemics (2009/10, 2020/21 and 2021/22). The model takes into account both the stochastic variability of the incidence and the uncertainty in the estimate of the reproduction number.

***Statistical-1*.** Linear autoregressive exogenous model in which the forecasts are based on past official data and an exogenous variable. The traditional surveillance data comes from the official national institute. The exogenous variable comes from the information retrieved from Influweb, a web-based participatory surveillance platform that has been monitoring ILI incidence in Italy since 2008 as part of the Influenzanet network [6, 7]. Influweb's participatory system offers the advantage of providing real-time data, a feature not present with traditional surveillance, which only offers data for the preceding week. Leveraging this real-time information, this model incorporates the instantaneous ILI signal from Influweb to forecast the upcoming four weeks. The approach involves generating predictions for ILI by integrating the ILI incidence official data from the three preceding weeks and the current week's signal from Influweb, averaged with the two preceding weeks, into a linear autoregressive exogenous model. The regression coefficients of the autoregressive model are estimated separately for the different time horizons, using a least squares regression.

***Statistical-2*.** Autoregressive integrated moving average (ARIMA) model trained by using data from the previous (2022-23) and current (2023- 24) influenza season.
